# Psychometric properties of the Barkin Index of Maternal Functioning with Ethiopian mothers

**DOI:** 10.1101/2021.12.01.21266886

**Authors:** Selamawit Gebeyehu, Yordanos Gizachew, Yilma Chisa, Beemnt Tekabe, Robert Trevethan

**Affiliations:** School of Public Health, College of Medicine and Health Science, Arba Minch University, Arba Minch, Gamo Zone, Ethiopia; School of Nursing, College of Medicine and Health Science, Arba Minch University, Arba Minch, Gamo Zone, Ethiopia; Independent academic researcher and author, Albury, NSW, Australia

**Author notes:** Corresponding Author: Selamawit Gebeyehu.

**Keywords:** Barkin Index of Maternal Functioning, BIMF, maternal functioning, Ethiopia

## Abstract

**Background:** The functional status of mothers after childbirth has implications for maternal, child, and family health. There is a lack of adequate reliable and valid instruments in Ethiopia for assessing women’s postpartum functional status.

**Objective:** This study was intended to reveal the psychometric properties of the Barkin Index of Maternal Functioning (BIMF) for assessing Ethiopian mothers’ functional status.

**Method:** Structured interviews were used to obtain BIMF data from 202 women who had a child less than 1 year of age. Descriptive statistics were calculated for the BIMF items; internal consistency was assessed with interitem correlations and coefficient alphas; construct validity was examined through exploratory factor analyses (EFAs) after face and content validities had been confirmed; and test–retest reliability was assessed with intraclass correlation coefficients (ICCs).

**Results:** Narrow standard deviations and significant skewness and kurtosis characterized most of the individual BIMF items. Most interitem correlations were < |.15| and 13 of the 20 BIMF items did not load satisfactorily on any factor in exploratory factor analyses. Two factors emerged from the remaining items, one with three items and the other with four. Coefficient alphas were .54 for the first of these factors, .48 for the second, and .58 for all 20 items. The ICCs for test– retest reliability were < .40.

**Conclusions:** BIMF data from the sample of Ethiopian women in this study exhibited unusually low levels of variability and high levels of skewness and kurtosis. Furthermore, in the context of this study, the BIMF could not be regarded as reliable either in terms of internal (interitem) consistency or temporal (test–retest) consistency. Researchers using the BIMF in Ethiopian contexts are advised to examine the nature of their data carefully, identify the factor structure of their samples’ data, and consider ways in which the index might be improved, or replaced.

## Introduction

Maternal functioning in the postpartum period is a reflection of the physical and psychosocial changes associated with pregnancy and childbirth. Maternal activities, in addition to physiological changes as well as individual and social activities, can make this period difficult for mothers. As a result, assessing maternal functioning following childbirth can be important for identifying maternal performance, productivity, and health, and can provide valuable information for healthcare providers, particularly with regard to identifying mothers who might benefit from supportive advice or interventions [1–3].

Almost all of the postpartum functional status assessment tools available today were developed for populations in high-income countries [4]. To date, the Inventory of Functional Status After Childbirth (IFSAC) is the most widely used tool specifically designed to assess several domains of maternal functioning [5]. The IFSAC contains 36 items arranged in subscales encompassing five dimensions: infant care responsibilities, self-care activities, household activities, social and community activities, and occupational activities. The inventory is designed to assess maternal functioning from 6 to 10 weeks after childbirth with a focus on “recovery from childbirth” [6]. Despite its popularity, the IFSAC has been criticized because it does not account for women’s feelings or levels of satisfaction with the changes in their lives since childbirth, and, as an alternative, the Barkin Index of Maternal Functioning (BIMF) was recommended by its developers as a more modern, patient-centered method for assessing maternal functioning [7, 8]. It is therefore the focus of this study.

The BIMF is a 20-item Likert-type scale designed to capture mothers’ perceptions of their functional status across the previous 2-week period at some point within the year since the birth of their most-recent, or only, child. For each item, participants are offered six response options: 0 corresponding to “strongly disagree”, 1 to “disagree”, 2 to “slightly disagree”, 3 to “neutral”, 4 to “slightly agree”, 5 to “agree”, and 6 to “strongly agree”. Items 16 and 18 are negatively worded, so responses on those items are reverse coded prior to analyzing the data to ensure that higher scores on the index consistently indicate higher levels of functoning.

Because of the lack of a valid means for assessing maternal functional status in sub-Saharan countries, including Ethiopia, in this article we examine the validity and reliability of the BIMF when used with Ethiopian mothers to identify whether the index is likely to perform satisfactorily within that population.

## Method

### Study design and setting

A descriptive community-based study was implemented with participants from Lante District within Gamo Zone, Ethiopia, situated 430 km south of the country’s capital, Addis Ababa. In 2020 G.C. (2012 E.C.), the district health office reported a total population of 236,600, with 436 mothers having a child under 1 year of age.

### Participants

Eligible participants were women who had a child less than 1 year of age resulting from a single-child pregnancy; who had vaginal delivery, birth through caesarean section, or instrumental delivery; who had self-declared physical health; and who had self-declared absence of depression or any other mental disorder during the pre-pregnancy, prepartum, and postpartum periods.

### Sample size determination and sampling method

Sample size required to determine the factor structure of the BIMF was based on there being at least 10 participants per item of the index [9]. Therefore, a sample of at least 200 mothers was required. In order to attain that sample, a list of the 436 mothers who had a child less than 1 year of age was obtained from the district health office. Subsequently, a sample of 202 mothers was identified by using computer-generated random numbers with Microsoft Excel for Windows 10. Forty of the 202 mothers were selected by the same method to comprise a retest sample for the BIMF, 2 weeks after the index had initially been administered.

### Questionnaire

A questionnaire was created that comprised questions to obtain information about the mothers’ sociodemographic background (the respondent and her most-recent child’s ages, the respondent’s marital status, and size of her family), obstetric characteristics (parity, number of living children, mode of delivery of the most recent child, and whether or not the mother had intended to become pregnant), and the BIMF.

### Translation process

Translation and cultural adaptation of the BIMF (for example, the word “diaper” is not familiar to most people in rural communities, and in Amharic is referred to as “ye shint cherq”) was performed according to the minimal translation criteria of the Medical Outcomes Trust [10]. Two independent bilingual translators (one a psychologist and the other an English-language lecturer) with advanced levels of English language and native Amharic-language skills translated the BIMF into Amharic. In doing so, they made no changes to the items’ meanings. An English-language lecturer who is a native Amharic speaker and who had been blinded to the original version, provided a back translation. There were no major difficulties in reconciling the back-translated version. Subsequently, Dr Barkin, the main researcher associated with development of the BIMF, gave permission for the Amharic version of the index to be used in this research.

### Face and content validity

After translation, the BIMF was reviewed by experts in maternity and child health and reproductive health units who were not part of the study, and they identified no problems. The tool was regarded as likely to be generally well understood, acceptable, and culturally appropriate for all prospective respondents.

### Data collection

Names and addresses of the 202 mothers identified as prospective participants were obtained from the district health office. Five data collectors and three field supervisors who had experience in conducting interviewer-administered data-collection procedures and the ability to speak Amharic were recruited for data collection. The data collectors went to the mothers’ homes and conducted structured interviews by using the smart-phone based Open Data Kit (ODK) app. Prospective participants were provided verbal and written information about the research, and were invited to ask any questions they might have. All mothers who were invited to participate in the research agreed to do so.

### Data processing and analysis

Data were analyzed using SPSS for Windows version 25^®^ (IBM Corp., Armonk, NY, USA). Descriptive statistics were calculated for the sociodemographic variables and BIMF items. Reliability associated with internal consistency was measured by interitem correlations and coefficient alphas.

Because previous research with the BIMF had revealed marked inconsistency regarding number and composition of factors in the scale [11–14], we did not conduct confirmatory factor analysis as a means of identifying the structure of the data. Instead, we conducted a series of exploratory factor analyses (EFAs). In doing so, we identified the number of factors with the scree test in conjunction with parallel analysis based on principal components and 1,000 randomly generated matrices; we used principal axis factoring as the method of extraction in conjunction with promax rotations; and we regarded item loadings > |0.40| as definitely acceptable, and loadings in the region of |.35| to |.40| as possibly acceptable.

In order to assess test–retest reliability, ICCs (3,1 [two-way mixed effects, single measures] absolute agreement) were calculated.

### Ethical considerations

Ethics approval was obtained from Arba Minch University’s College of Medicine and Health Science Institutional Review Board (IRB/157/2020). Verbal consent was obtained from all participants and recorded in conjunction with the starting time of each interview, and all interviews were conducted in private.

## Results

### Participant characteristics

All 202 mothers reported their physical health to be good, and none indicated they had a history of depression or any other mental disorder during the pre-pregnancy, prepartum, and postpartum periods. Their ages ranged from 15 to 38 years (*M* = 26.1 years, mode = 30, *SD* = 4.7), their children’s ages ranged from 1 to 12 months (*M* = 6.7 months, mode = 11, *SD* = 3.7), their number of pregnancies ranged from 1 to 9 (*M* = 2.4, mode = 1, *SD* = 1.5), their number of children ranged from 1 to 8 (*M* = 2.31, mode = 1, *SD* = 1.40), and the size of their households ranged from 2 to 12 (*M* = 4.6, mode = 4, *SD* = 1.7). Other participant characteristics are shown in Table 1.

**Table 1.** Participant characteristics.

| Variable | Category | Number | Percent |
| --- | --- | --- | --- |
| Age of the mother/<br>caregiver (in years) | $\leq 24$ | 73 | 36.1 |
|  | 25 – 30 | 103 | 51.0 |
| | $\geq 31$ | 26 | 12.9 |
| Age of most recent or<br>only child | < 2 months | 20 | 9.9 |
|  | 2 – 6 months | 76 | 37.6 |
| | $\geq 6$ months | 106 | 52.6 |
| Marital status | Single | 9 | 4.5 |
|  | Married | 191 | 94.6 |
|  | Divorced or<br>widowed | 2 | 1 |
| Family size | $\leq 3$ | 58 | 28.7 |
|  | 4 – 6 | 115 | 56.9 |
| | $\geq 7$ | 29 | 14.4 |
| **Parity** | Primipara | 69 | 34.2 |
|  | Multipara | 133 | 65.8 |
| **Number of living children** | $\leq 3$ | 165 | 81.7 |
| | $\geq 4$ | 37 | 18.3 |
| **Mode of delivery** | Vaginal | 164 | 81.2 |
|  | Caesarean | 25 | 12.4 |
|  | Instrument | 13 | 6.4 |
| **Pregnancy intention** | Intended | 175 | 86.6 |
|  | Unintended | 27 | 13.4 |

### Item characteristics

Means, *SD*s, and indicators of skewness and kurtosis on the 20 BIMF items at Time 1 are provided in Table 2. On all but one of the items, the means were above the midpoint of the option range, indicating a tendency among participants to believe they were functioning effectively as mothers. The exception was Item 18, on which, prior to reverse coding, the participants indicated they were often anxious or worried about fulfilling their maternal responsibilities. Twelve of the items had *SD*s < 1.0; 18 items were significantly skewed (two positively, 16 negatively); and 15 items were significantly kurtotic (12 leptokurtic, three platykurtic). Twelve items were significantly negatively skewed as well as being significantly leptokurtic.

**Table 2.**
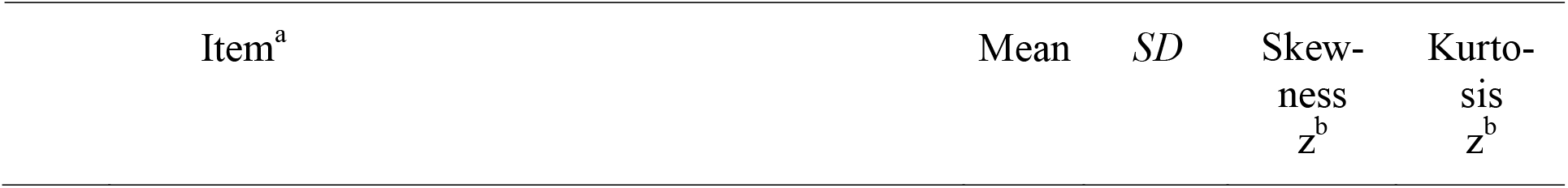
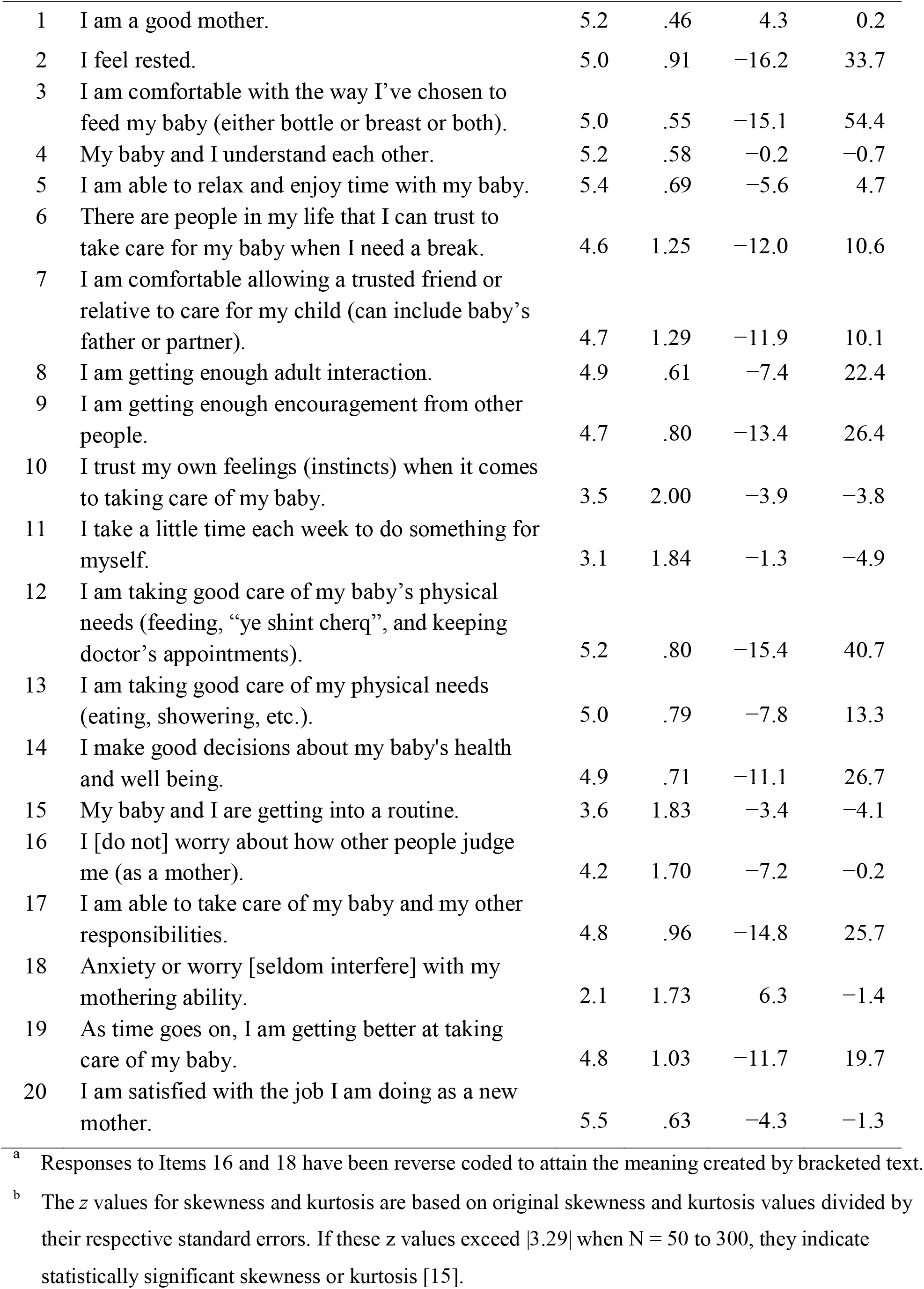
Descriptive statistics for items on the Barkin Index of Maternal Functioning.

| Item<sup>a</sup> | Mean | *SD* | Skew-<br>ness<br>*z*<sup>b</sup> | Kurto-<br>sis<br>*z*<sup>b</sup> |
| --- | --- | --- | --- | --- |

|  |  |  |  |  |  |
| --- | --- | --- | --- | --- | --- |
| 1 | I am a good mother. | 5.2 | .46 | 4.3 | 0.2 |
| 2 | I feel rested. | 5.0 | .91 | -16.2 | 33.7 |
| 3 | I am comfortable with the way I've chosen to feed my baby (either bottle or breast or both). | 5.0 | .55 | -15.1 | 54.4 |
| 4 | My baby and I understand each other. | 5.2 | .58 | -0.2 | -0.7 |
| 5 | I am able to relax and enjoy time with my baby. | 5.4 | .69 | -5.6 | 4.7 |
| 6 | There are people in my life that I can trust to take care for my baby when I need a break. | 4.6 | 1.25 | -12.0 | 10.6 |
| 7 | I am comfortable allowing a trusted friend or relative to care for my child (can include baby's father or partner). | 4.7 | 1.29 | -11.9 | 10.1 |
| 8 | I am getting enough adult interaction. | 4.9 | .61 | -7.4 | 22.4 |
| 9 | I am getting enough encouragement from other people. | 4.7 | .80 | -13.4 | 26.4 |
| 10 | I trust my own feelings (instincts) when it comes to taking care of my baby. | 3.5 | 2.00 | -3.9 | -3.8 |
| 11 | I take a little time each week to do something for myself. | 3.1 | 1.84 | -1.3 | -4.9 |
| 12 | I am taking good care of my baby's physical needs (feeding, "ye shint cherq", and keeping doctor's appointments). | 5.2 | .80 | -15.4 | 40.7 |
| 13 | I am taking good care of my physical needs (eating, showering, etc.). | 5.0 | .79 | -7.8 | 13.3 |
| 14 | I make good decisions about my baby's health and well being. | 4.9 | .71 | -11.1 | 26.7 |
| 15 | My baby and I are getting into a routine. | 3.6 | 1.83 | -3.4 | -4.1 |
| 16 | I [do not] worry about how other people judge me (as a mother). | 4.2 | 1.70 | -7.2 | -0.2 |
| 17 | I am able to take care of my baby and my other responsibilities. | 4.8 | .96 | -14.8 | 25.7 |
| 18 | Anxiety or worry [seldom interfere] with my mothering ability. | 2.1 | 1.73 | 6.3 | -1.4 |
| 19 | As time goes on, I am getting better at taking care of my baby. | 4.8 | 1.03 | -11.7 | 19.7 |
| 20 | I am satisfied with the job I am doing as a new mother. | 5.5 | .63 | -4.3 | -1.3 |

<sup>a</sup> Responses to Items 16 and 18 have been reverse coded to attain the meaning created by bracketed text.
<sup>b</sup> The $z$ values for skewness and kurtosis are based on original skewness and kurtosis values divided by their respective standard errors. If these $z$ values exceed $|3.29|$ when $N = 50$ to $300$ , they indicate statistically significant skewness or kurtosis [15].

### Interitem correlations and factor structure

None of the 190 interitem correlations exceeded .50, and only 37% of those correlations lay in the .15 to .50 range recommended by Clark and Watson [16]. Among the remaining 63% of the interitem correlations, 17% were negative, with the strongest being −.29. Despite this generally low, and sometimes inverse, level of association between the items, the Kaiser–Meyer–Olkin (KMO) index was .72 and Bartlett’s test of sphericity was significant (*p* < .001), and therefore the data appeared to be suitable for EFA. Among the 20 items, seven eigenvalues were > 1, but the scree plot indicated there were four factors in the data (see Fig 1), as did parallel analysis. We therefore conducted an initial EFA constrained to four factors. In this solution, nine items loaded uniquely on one of the factors at > |.40|, three items had cross loadings on two of the factors, and the remaining eight items failed to load on any factor at > |.37|. Only 42.56% of the variance was accounted for by the four factors.

[Fig1. Scree plot based on all 20 BIMF items.]

Follow-up EFAs were conducted in order to attain the most satisfactory and interpretable outcome. When nonloading items, cross-loading items, and items that loaded only on single-item factors were successively removed, seven items were retained, among which 53.09% of the variance was accounted for, but the extraction communalities were generally low (maximum = .55, mean = .35, with Items 20 and 18 exceptionally low at .23 and .13, respectively). All seven items loaded at > .39 on one of two factors, although two items cross loaded with a difference ≤ .20. See Table 3. Items 13, 14, and 17 loaded on a factor that we labeled Decision Making and Practicalities, and Items 5, 16, 18, and 20 loaded on the other factor, which we labeled Confidence, Enjoyment, and Satisfaction.

**Table 3.** Item loadings on the two-factor solution.^a^.

| Item # | Item<sup>b</sup> | Factor 1 | Factor 2 |
| --- | --- | --- | --- |
| 14 | I make good decisions about my baby's health and well-being. | .74 |  |
| 17 | I am able to take care of my baby and my other responsibilities. | .56 | -.40 |
| 13 | I am taking good care of my physical needs (eating, showering, etc.). | .52 |  |
| 16 | I [do not] worry about how other people judge me (as a mother). |  | .64 |
| 5 | I am able to relax and enjoy time with my baby. | .32 | .52 |
| 20 | I am satisfied with the job I am doing as a new mother. |  | .42 |
| 18 | Anxiety or worry [seldom interfere] with my mothering ability. |  | .39 |

<sup>a</sup> Loadings $< |.30|$ are not shown.
<sup>b</sup> Wording of Items 16 and 18 was altered, as shown in bracketed text, to reflect prior reverse coding.

Consonant with the practice of several other researchers [11, 13, 14], we calculated a score for each subscale by adding responses on their respective items, and we calculated a composite total by adding responses to all 20 items. In order to confer meaning to participants’ responses in relation to the range of response options, we also divided these totals by the number of items on which each score was based. The results are shown in Table 4.

**Table 4.** Means, standard deviations, and obtainable ranges on composite factor and total scores.^a^.

| BIMF<sup>b</sup> component and items | Unadjusted scores |  | Adjusted scores |  |
| --- | --- | --- | --- | --- |
|  | Mean (*SD*) | Obtainable range | Mean (*SD*) | Obtainable range |
| Decision Making and Practicalities<br>(Items 13, 14, 17) | 14.74 (1.79) | 0–18 | 4.91 (0.60) | 0–6 |
| Confidence, Enjoyment, and<br>Satisfaction (Items 5, 16, 18, 20) | 17.08 (1.79) | 0–24 | 4.27 (0.81) | 0–6 |
| Total (Items 1 to 20) | 91.11 (7.76) | 0–120 | 4.56 (0.39) | 0–6 |

<sup>a</sup> Unadjusted scores are based on addition of mothers' responses; adjusted scores were obtained by dividing the unadjusted scores by the relevant number of items.
<sup>b</sup> Barkin Index of Maternal Functioning

The means of the adjusted scores on both factors as well as the composite total indicate that the mothers tended to respond with options of somewhat agree (4) or agree (5). However, relative to the first factor (Decision Making and Practicalities), the means and *SD*s indicated that self-perceptions of maternal functioning were lower and more varied on the second factor (Confidence, Enjoyment, and Satisfaction). Scores on the first factor were significantly skewed (−11.76) and kurtotic (21.9), and the total score was significantly skewed (−3.4).

Coefficient alphas were .54 for Decision Making and Practicalities, .48 for Confidence, Enjoyment, and Satisfaction, and .58 for all 20 items.

### Test–retest results

The ICC for Confidence, Enjoyment, and Satisfaction could not be calculated satisfactorily because of negative average covariance among the items (Pearson’s *r* of −.04 was calculated as a substitute for the ICC). The ICC for Decision Making and Practicalities was .14 [95% CI −.18– .43], and the ICC for the total score across all 20 items was .36 [95% CI .06–61].

## Discussion

To the authors’ best knowledge, this is the first examination of the BIMF in an Ethiopian context. The research has several desirable foundations. Face and content validity of the BIMF were assessed and regarded as satisfactory; random sampling was used to obtain participants; the sample included women who gave birth by vaginal delivery, instrument, and caesarean section, and who were primiparous as well as multiparous, suggesting that the results were not likely to be unique to a specific group of mothers; the sample was sufficiently large for identifying the factor structure of the data; and an attempt was made to identify test–retest reliability.

Three aspects of the raw data are noteworthy and might carry implications for assessing the suitability of the BIMF in Ethiopian settings. First, with the exception of Item 18 (referring to mothers’ worry or anxiety), the participants’ responses on individual items suggest a moderate to high level of maternal functioning. Whether that is an accurate indication would require independent assessment, which was not conducted in this research. However, some reassurance might be taken from the mean of 91 on the 20 BIMF items in this research being similar to the mean of the 20 items in research where samples were composed solely of mothers who were not depressed [13, 17]. Furthermore, in samples that include a large proportion of mothers diagnosed with depression and who could therefore be expected to have lower 20-item BIMF scores, the mean based on those items has indeed been lower, for example, ranging from 81 down to 63 [7, 11, 17, 18]. However, in a sample of mothers, 46% of whom had been assessed as experiencing depression, the 20-item BIMF score was considerably higher, at 104 [19]. Unfortunately, this range of 40 points on the total BIMF score in samples with mothers who experience depression makes it difficult to place confidence in the meaning of the 20-item BIMF score when seeking to gain a definitive overview of mothers’ functioning, and the composite score of 91 in this research therefore provides little reassurance in relation to validity of the BIMF in an Ethiopian setting.

The second noteworthy aspect of the data is the lack of variability among the mothers’ responses. This was evident in 60% of the *SD*s on individual items being < 1.00 despite the availability of seven response options. In contrast, only two of the *SD*s were < 1 on the 20 items in the foundational psychometric research with the BIMF [11]. Furthermore, the *SD* of the composite score based on all 20 items in this research was 7.76 compared with noticeably wider *SD*s, ranging from 10.8 in some research [20] up to 17.1 in other research [7].

This smaller amount of variation in the mothers’ responses in the present study presents interpretative and empirical difficulties for three reasons. First, if there is genuine lack of variability, statistics such as correlations, factor analysis, and coefficient alpha—all of which rely on variability—are likely to yield misleading results. Second, if the lack of variability has a foundation in some kind of distorting BIMF-related artifact within Ethiopian samples (e.g., the cognitive burden on the mothers having to remember what occurred during the previous 2 weeks might have created a degree of bluntness in their responses), the nature of that artifact would need to be identified and addressed. Third, if the lack of variability is likely to be a chance occurrence, other studies need to be conducted with the prospect that their data would contain a greater spread of maternal self-impressions.

The third noteworthy aspect of the data is most items having distributions that were significantly skewed or kurtotic, or both, which was accompanied by one of the factors and the 20-item composite score having negatively skewed distributions. These results could be genuine reflections of the mothers’ self-perceptions, and therefore are interesting findings in themselves. However, in combination with the minimal variation in responses to the 20 items, the skewed and kurtotic distributions might also have influenced the EFA outcome, coefficient alpha values, and the test–retest results.

We are aware of no other research in which a similar BIMF factor structure emerged. Although two factors have been found in some other studies [11, 14], the factors in those studies comprised different items and were therefore conceived of as being different and were labeled differently. Our results are also distinctive in that 13 of the 20 BIMF items failed to load on any factor, probably because most interitem correlations were < |.15|. An additional feature of our results is Items 16 and 18 having loaded on one of our factors (Confidence, Enjoyment, and Satisfaction), but having loaded on none of the factors in some other research [11, 12, 14].

Although inconsistencies in factor structure could result from differences in the nature of samples, particularly when cultural differences exist, some inconsistency between our findings and the findings in other research is likely to result from principal components analysis versus EFA being employed as a data-reduction strategy; different procedures (the Kaiser criterion, scree plots, or parallel analysis) being used for identifying the number of factors in the data; the method used for extraction (e.g., maximum likelihood or principal axis factoring); whether or not the factors were rotated; the method employed for rotation (orthogonal versus oblique)—if factor rotation occurred at all; and whether or not a sequence of EFAs was used to attain greater factor clarity by successively deleting poorly performing items. Publications about the BIMF provide little or no information about these procedures when data-reduction analyses have been conducted despite some procedures having substantial implications for the outcomes.

In this research, we believe we engaged in best-practice data-reduction procedures [21–26], so the results are likely to have been dependent on the nature of the data, not deficiencies in the method of analysis. That aside, the EFA results in this research are not impressive. Not only were loadings on 13 of the 20 BIMF items so low that most of the index’s items were discarded, but also the remaining seven items had low communalities despite accounting for 53.1% of the variance. Furthermore, two of the seven items cross loaded on the two factors that emerged. Under these circumstances, one solution involves focusing on the composite score based on all 20 items, but mothers with similar scores could have noticeably different profiles and therefore noticeably different needs in relation to healthcare as well as correlates in research contexts, so the value of that score is limited.

In addition to differences in factor structure, our results differ in several ways from the results obtained in other studies. The coefficient alpha across all 20 items in our data was unusually low at .58 compared with alphas of .87 and .83 in samples of American mothers [7, 11, 19] and, at .36, the test–retest ICC for the 20-item composite score in this research indicated much poorer test–retest reliability than was indicated by the ICC of .80 obtained in a study conducted with Iranian mothers [14]. The above differences, in which the psychometric features of the BIMF in this research are not only less impressive than are results reported in other BIMF research but are also unimpressive according to conventional criteria, are almost definitely attributable to limited data variability in this study [27, 28].

Although not directly associated with our examination of the BIMF, the absence of self-reported depression among the mothers in this research might be important. Other research has indicated that perinatal depression is likely to be approximately 13% among mothers in middle- and low-income countries in general [29] and in Ethiopia, specifically [30], and as high as 33% in rural parts of Ethiopia [31]. The discrepancy between those statistics and the mothers’ self-reports in this research suggests that responses on the BIMF in this research might be regarded with a degree of reservation despite the mothers having been assured by the data collectors of confidentiality concerning their responses in order to discourage them from creating an inaccurately favorable impression.

## Conclusions

This study provides ambiguous evidence concerning the reliability and validity of the BIMF in an Ethiopian setting, largely because it is difficult to identify whether the generally high and similar self-perceptions of mothers were genuine and might also have been idiosyncratic to our specific sample.

The ambiguity could lead to incorrect conclusions that, in Ethiopian samples, the BIMF’s items carry little association with each other; that there are only two factors within the BIMF, each with only a small number of items; and that the index’s test–retest reliability is poor. Because these conclusions might be proved untenable if data were obtained from other samples, including samples with a greater dispersion among responses, the BIMF may be appropriate in other Ethiopian contexts. Furthermore, with more variation in the data there is a prospect that additional, or more substantial, factors would emerge.

Because the two factors that emerged from the EFAs were dissimilar to factors found in other research, and because of evident volatility in the factor structure of the BIMF revealed by other studies, further use of the BIMF in an Ethiopian context should involve examination of the factor structure for the particular sample(s) on which that research is conducted. Furthermore, because our research indicates that the index could not be regarded as reliable either in terms of internal (interitem) consistency or temporal (test–retest) consistency, prospects for improving the index might be considered—although with the proviso that permission to modify the index had been granted [32].

The seven BIMF items that loaded on two factors in this research might be used to explore aspects of decision making and practicalities, as well as aspects of enjoyment and satisfaction, among Ethiopian women concerning their maternal role in the postpartum period. However, given the volatility of the BIMF factor structure, those two factors might not feature in data from other samples. Furthermore, if only those seven items were used, important aspects of motherhood such as mother–infant interaction, self-care, and social support would not be assessed, and this raises the possibility that, when face and content validity were assessed at the start of this research, too much consideration was given to endorsing the items within the BIMF without considering aspects of motherhood that were underrepresented, inappropriately represented, or not represented at all — at least for an Ethiopian context.

In summary, this research demonstrates that the BIMF should be used with a degree of caution in Ethiopian contexts and that, if researchers want to use it, they should anticipate obtaining sufficiently large samples to be able to conduct EFAs to identify the factor structure of the data produced by those samples and also consider ways in which the index might be modified if doing so is likely to result in more varied responses — with the underlying proviso that the greater variability was associated with genuine differences in self-perception. An alternative course of action would be to seek, or create, instruments that provide greater applicability and usefulness for assessing maternal functioning in Ethiopian contexts.

## Data Availability

All data produced in the present study are available upon reasonable request to the authors

## Acknowledgements

The authors thank the participants for their valuable information and the data collectors and supervisors for their commitment to this research. We also extend our gratitude to Arba Minch University for support and opportunities provided for us to conduct this study.

## Abbreviations and acronyms

BIMF: Barkin Index of Maternal Functioning
EFA: exploratory factor analysis
ICC: intraclass correlation coefficient
IFSAC: Inventory of Functional Status After Childbirth

